# Impact of vaccination on post-acute sequelae of SARS CoV-2 infection in patients with rheumatic diseases

**DOI:** 10.1101/2022.10.06.22280798

**Authors:** Naomi J. Patel, Claire Cook, Kathleen M.M. Vanni, Xiaoqing Fu, Xiaosong Wang, Yumeko Kawano, Grace Qian, Buuthien Hang, Shruthi Srivatsan, Emily Banasiak, Emily Kowalski, Katarina Bade, Yuqing Zhang, Jeffrey A. Sparks, Zachary S. Wallace

## Abstract

**Objective:** Vaccination decreases the risk of severe COVID-19 but its impact on post-acute sequelae of COVID-19 (PASC) is unclear among patients with systemic autoimmune rheumatic diseases (SARDs) who may have blunted vaccine immunogenicity and be vulnerable to PASC.

**Methods:** We prospectively enrolled SARD patients from a large healthcare system who survived acute infection to complete surveys. The symptom-free duration and the odds of PASC (any symptom lasting ≥ 28 or 90 days) were evaluated using restricted mean survival time and multivariable logistic regression, respectively, among those with and without breakthrough infection (≥ 14 days after initial vaccine series).

**Results:** Among 280 patients, the mean age was 53 years, 80% were female, and 82% were white. The most common SARDs were inflammatory arthritis (59%) and connective tissue disease (24%). Those with breakthrough infection had more upper respiratory symptoms, and those with non-breakthrough infection had more anosmia, dysgeusia, and joint pain. Compared to those with non-breakthrough COVID-19 infection (n=164), those with breakthrough infection (n=116) had significantly more symptom-free days over the follow-up period (+28.9 days, 95% CI: 8.83, 48.89; p=0.005) and lower odds of PASC at 28 and 90 days (aOR 0.49, 95% CI: 0.29, 0.83 and aOR 0.10, 95% CI: 0.04, 0.22, respectively).

**Conclusion:** Vaccinated patients with SARDs were less likely to experience PASC compared to those not fully vaccinated. These findings support the benefits of vaccination for patients with SARDs and suggest that the immune response to acute infection is important in the pathogenesis of PASC in SARD patients.

**Key Messages:** *What is already known on this topic?:*

- Post-acute sequelae of COVID-19 (PASC) affects 20-50% of COVID-19 survivors, though the impact of vaccination on the risk and severity of PASC is unclear, especially among those with systemic autoimmune rheumatic diseases (SARDs) who may have impaired responses to vaccines and be particularly vulnerable to PASC.

*What this study adds?:*

- In this prospective cohort of SARD patients recovering from COVID-19, we found that those with breakthrough vs non-breakthrough infection had more symptom-free days over the follow-up period (adjusted difference +28.9 days, 95% CI: 8.38, 48.89; p=0.005) and a lower odds of PASC at 28 days (aOR 0.49, 95% CI: 0.29, 0.83) and at 90 days (aOR 0.10, 95% CI: 0.04, 0.22).
- Patient-reported pain and fatigue scores were lower, reflecting less severe pain and fatigue, in those with breakthrough infection compared to those with non-breakthrough infection.

*How this study might affect research, practice, or policy?:*

- This study extends our understanding of the benefits of vaccination against COVID-19 in patients living with SARDs and reinforces the importance of vaccinating this vulnerable population.
- Our findings suggest that the initial immune response to acute SARS-CoV-2, as influenced by vaccination, affects PASC risk but this requires further study.

## Introduction

Patients with systemic autoimmune rheumatic diseases (SARDs) are at higher risk of severe acute outcomes of COVID-19 infection, though few studies have investigated the risk of longer-term complications of COVID-19.^1-7^ Vaccines are safe and efficacious in reducing risk for severe COVID-19 among SARD patients, but less is known about how they may impact the risk of post-acute sequelae of COVID-19 (PASC).

PASC refers to persistent symptoms following acute infection and is currently defined by duration of symptoms, with some requiring symptoms that persist for at least one month and others for at least three months following acute infection.^8,9^ PASC incorporates a heterogeneous set of symptoms that may include impaired executive function, fatigue, dyspnea, cough, palpitations, myalgias or arthralgias, and/or anosmia, among others. A higher severity of acute COVID-19 is associated with a greater risk of PASC, though asymptomatic patients or those with minimal symptoms can also develop PASC.^1,10,11^ Population-based studies suggest that PASC can affect between 20-40% of people, and up to 50-70% of hospitalized patients may continue to have symptoms even months following hospital discharge.^10-13^ Patients with SARDs may be vulnerable to PASC due to altered immunity, immunosuppressive therapy, and increased risk for severe acute COVID-19.

While vaccination against SARS-CoV-2 decreases the risk of severe acute outcomes, there are limited data regarding the effect of vaccination on PASC risk.^14,15^ Previous studies of patients without SARDs have suggested a decreased risk of PASC in those who were vaccinated prior to COVID-19 infection.^16-18^ However, many patients with SARDs have impaired responses to the SARS-CoV-2 vaccine and may not similarly benefit from vaccination with regard to the risk of PASC.^19-23^ In this study, we investigated the association of SARS-CoV-2 vaccination with the risk of PASC in patients with SARDs.

## Methods

### Study population and patient identification

We performed a prospective study in Mass General Brigham (MGB), a large, multi-center healthcare system that includes two tertiary care hospitals (Massachusetts General Hospital and Brigham and Women’s Hospital), twelve community hospitals, and their associated primary and specialty outpatient centers in the greater Boston, Massachusetts, area. We identified patients within MGB who were ≥18 years of age, had a positive test result for SARS-CoV-2 by polymerase chain reaction (PCR) or antigen nasopharyngeal test between March 1, 2020 and July 8, 2022, and had a rheumatic disease diagnosis based on billing codes. Diagnosis of a prevalent SARD at the time of infection was then confirmed by manual review of the EHR. This approach has been previously described.^2,6,7^ This study was approved by the MGB Institutional Review Board (2020P000833).

### Patient recruitment for prospective study

From this population, we invited patients who survived their acute infection to participate in a prospective, longitudinal study: COVID-19 and Rheumatic Diseases (RheumCARD). As previously described in detail, potential participants were invited to participate either via secure online EHR portal or US mail.^2^ Initial invitations were sent on March 11, 2021, and as new subjects with confirmed infection were identified, subsequent patients were invited on a rolling basis, approximately once per month, at least 28 days following their COVID-19 diagnosis date.

### Data Collection

Demographic data assessed in the survey included age, sex, race, and ethnicity. Smoking status was assessed as never, past, or current. The comorbidity count was derived as the sum of comorbidities queried. COVID-19 symptoms assessed in the survey included fever, sore throat, new cough, nasal congestion/rhinorrhea, dyspnea, chest pain, rash, myalgia, fatigue/malaise, headache, nausea/vomiting, diarrhea, anosmia, dysgeusia, and joint pain. The symptom count was calculated as the sum of these self-reported symptoms. We collected details of the acute COVID-19 course, including symptom duration, treatments, and details of hospitalization (if applicable). Time to COVID-19 symptom resolution and vaccination status were collected.

SARDs were categorized broadly as inflammatory arthritis (including rheumatoid arthritis, psoriatic arthritis, juvenile idiopathic arthritis, axial spondyloarthropathy, or other inflammatory arthritis), vasculitis (including ANCA-associated vasculitis, giant cell arteritis and/or polymyalgia rheumatica, or other vasculitis such as Takayasu arteritis or Kawasaki disease), connective tissue disease (CTD, including systemic lupus erythematosus, mixed connective tissue disease, undifferentiated connective tissue disease, idiopathic inflammatory myopathy, or Sjogren’s syndrome), or other (sarcoidosis, Behcet disease, IgG4-related disease, or relapsing polychondritis). Use of immunomodulator medications at the time of acute COVID-19 infection was assessed.

### Exposure of Interest

The exposure of interest was being fully vaccinated at COVID-19 onset versus partially vaccinated or unvaccinated. Based on patient report, we classified patients as fully vaccinated at the index date (date of COVID-19 diagnosis) if infection was ≥14 days after completion of their primary vaccine series according to the US Centers for Disease Control and Prevention (CDC) definition: two doses of a messenger ribonucleic acid (mRNA) SARS-CoV-2 vaccine (i.e., either BNT162b2 [Pfizer-BioNTech] or mRNA-1273 [Moderna]) or one dose of the Ad26.COV2.S (Johnson & Johnson-Janssen) vaccine.^24^ Other patients were classified as either partially vaccinated or unvaccinated at the index date.

### Outcome Assessments

The primary outcome was PASC, defined as any persistent symptom at least 28 days post-COVID-19 infection (US CDC definition).^9^ A secondary outcome was PASC, as defined by any persistent symptom at least 90 days post-COVID-19 infection (World Health Organization [WHO] definition).^8^ All patients were enrolled at least 28 days after their COVID-19 diagnosis. Only those who completed the surveys at least 90 days following their COVID-19 diagnosis were included in the analysis of the WHO definition of PASC. Symptom duration (in days) and symptom-free days were other secondary outcomes; days of symptoms were counted from the index date through the time of symptom resolution or date of survey completion if symptoms were ongoing. Patients with missing data regarding symptom duration or vaccination status were excluded.

Other secondary outcomes included pain (measured by the short-form McGill Pain Questionnaire [SF-MPQ^25^]), fatigue (Fatigue Symptom Inventory [FSI^26,27^]), and functional status (modified Health Assessment Questionnaire [mHAQ^28^]). The 12-item short-form health survey (SF-12) was used as a general measure of both physical and mental health status.^29^ A Physical Component Summary Score (PCS-12) and Mental Component Summary Score (MCS-12) were calculated. Among those who developed PASC, we compared pain, fatigue, functional status, and overall health status scores between those with PASC following breakthrough versus non-breakthrough COVID-19 infection. We also assessed rheumatic disease activity following COVID-19 infection, based on self-reported SARD flare, participant global assessment, and disease activity, as assessed by the RAPID-3 score.

Outcomes were assessed from March 11, 2021 (the time of completion of the first survey) through August 8, 2022 (the time of completion of the last survey at the time of manuscript preparation).

### Statistical analysis

Categorical variables are presented as number (percentage), and continuous variables are presented as mean ± standard deviation or median ± interquartile range, as appropriate. Continuous variables were compared using a two-sample t-test for continuous normally distributed variables or Wilcoxon test for continuous non-normally distributed variables. Categorical variables were compared using Chi-square tests.

We calculated odds ratios for PASC at 28 and 90 days using unadjusted and multivariable adjusted logistic regression. The first multivariable model adjusted for age, sex, and race. The second multivariable model adjusted for age, sex, race, comorbidity count, and use of any one of the following medications: anti-CD20 monoclonal antibodies, methotrexate, mycophenolate, or glucocorticoids. These medications were chosen because of their impact on SARS-CoV-2 vaccine immunogenicity.

To assess the robustness of our findings, we conducted four sensitivity analyses evaluating odds ratios for PASC as well as differences in other patient-reported outcomes, limiting the population to: 1) those who did not receive nirmatrelvir/ritonavir or monoclonal antibodies, 2) those who did not receive any COVID-19-related treatment, 3) those who completed the questionnaires within 6 months of COVID-19 infection, and 4) those who did not require hospitalization for acute COVID-19 infection.

To assess differences in the symptom-free time between those with breakthrough versus non-breakthrough infection, we used restricted mean survival time (RMST).^30-32^ The event in this analysis was the number of days to COVID-19 symptom resolution. We compared the areas under the cumulative incidence curves, representing the number of days following symptom resolution (symptom-free days) using the Kaplan-Meier method, and applied inverse probability weighting method to adjust for age, sex, race, comorbidity count, and exposure to anti-CD20 monoclonal antibodies, methotrexate, mycophenolate, or glucocorticoids. The difference between the two groups reflects the difference in the number of symptom-free days between the two groups, with the non-breakthrough group as the reference group. RMST has multiple strengths, including no required assumptions regarding proportional hazards as well as ease of interpretation (effect estimate in difference in number of days over a specified time interval as opposed to a hazard ratio). Our primary follow-up period was 204 days, given that this was the maximum follow-up period among those with breakthrough infections. We performed secondary analyses assessing these outcomes at 28 and 90 days.

The level of significance was set as a two-tailed p<0.05, and statistical analyses were completed using SAS statistical software (version 9.4; SAS Institute, Inc.).

## Results

### Participant Characteristics

We analyzed 280 patients with SARDs who survived COVID-19. One hundred and sixteen (41%) had a breakthrough COVID-19 infection and the remainder (164, 59%) were either unvaccinated or were partially vaccinated at the time of diagnosis and were considered to have non-breakthrough COVID-19 infection. The breakthrough and non-breakthrough groups were similar with respect to age, sex, race, ethnicity, smoking status, and SARD category (**Table 1**).

**Table 1.** Demographics and rheumatic disease characteristics of patients with history of COVID-19 infection

| **Characteristic** | **All rheumatic disease patients (N=280)** | **Breakthrough COVID-19 infection (N=116)** | **Non-breakthrough COVID-19 infection (N=164)** | **p-value** |
| --- | --- | --- | --- | --- |
| Age at time of COVID-19 symptom onset in years, mean ± SD | 52 (15) | 53 (15) | 51 (16) | 0.48 |
| Age at time of survey in years, mean ± SD | 53 (15) | 53 (15) | 52 (16) | 0.68 |
| Female, n (%) | 223 (80) | 93 (80) | 130 (79) | 0.88 |
| Race, n (%) |  |  |  |  |
| Asian | 12 (4) | 5 (4) | 7 (4) | 1.00 |
| Black | 20 (7) | 4 (3) | 16 (10) | 0.06 |
| White | 230 (82) | 101 (87) | 129 (79) | 0.08 |
| Other | 15 (5) | 6 (5) | 9 (5) | 1.00 |
| Unknown | 7 (3) | 3 (3) | 4 (2) | 1.00 |
| Hispanic or Latinx ethnicity | 27 (10) | 8 (7) | 19 (12) | 0.15 |
| Smoking status |  |  |  | 0.65 |
| Current | 4 (1) | 2 (2) | 2 (1) |  |
| Former | 72 (26) | 27 (23) | 45 (27) |  |
| Never | 203 (73) | 87 (75) | 116 (71) |  |
| SARD category^ |  |  |  | 0.33 |
| Inflammatory arthritis | 165 (59) | 61 (53) | 104 (63) |  |
| Connective tissue disease | 68 (24) | 30 (26) | 38 (23) |  |
| Vasculitis | 26 (9) | 13 (11) | 13 (8) |  |
| Multiple | 9 (3) | 5 (4) | 4 (2) |  |
| Other | 12 (4) | 7 (6) | 5 (3) |  |
| Immunomodulatory medications |  |  |  |  |
| Conventional synthetic DMARDs |  |  |  |  |
| Antimalarial (includes hydroxychloroquine and chloroquine) | 64 (23) | 34 (29) | 30 (18) | 0.04 |
| Methotrexate | 60 (21) | 27 (23) | 33 (20) | 0.56 |
| Mycophenolate mofetil/mycophenolic acid | 21 (8) | 14 (12) | 7 (4) | 0.02 |
| Other csDMARD* | 25 (9) | 14 (12) | 11 (7) | 0.14 |
| Targeted synthetic DMARD (JAK inhibitor) | 13 (5) | 7 (6) | 6 (4) | 0.40 |
| Biologic DMARDs |  |  |  |  |
| Anti-CD20 monoclonal antibody | 23 (8) | 14 (12) | 9 (5) | 0.08 |
| TNF inhibitor | 63 (22) | 28 (24) | 35 (21) | 0.66 |
| Other biologic DMARD<sup>†</sup> | 35 (13) | 15 (13) | 20 (12) | 0.86 |
| Baseline glucocorticoid use at COVID-19 onset | 51 (18) | 24 (21) | 27 (16) | 0.43 |
| Dose (prednisone-equivalent, daily mg), median [IQR] | 9 (5, 15) | 8 (5, 20) | 10 (5, 15) | 0.43 |
| Comorbidities |  |  |  |  |
| Obesity | 58 (21) | 17 (15) | 41 (25) | 0.04 |
| Hypertension | 64 (23) | 28 (24) | 36 (22) | 0.67 |
| Asthma | 49 (18) | 23 (20) | 26 (16) | 0.43 |
| Obstructive sleep apnea | 21 (8) | 7 (6) | 14 (9) | 0.50 |
| Coronary artery disease | 18 (6) | 6 (5) | 12 (7) | 0.62 |
| Diabetes | 16 (6) | 7 (6) | 9 (5) | 1.00 |
| Heart failure | 6 (2) | 2 (2) | 4 (2) | 1.00 |
| Chronic kidney disease | 12 (4) | 7 (6) | 5 (3) | 0.24 |
| Chronic obstructive pulmonary disease | 4 (1) | 2 (2) | 2 (1) | 1.00 |
| Interstitial lung disease/pulmonary fibrosis | 9 (3) | 5 (4) | 4 (2) | 0.50 |
| Solid tumor | 4 (1) | 1 (1) | 3 (2) | 0.64 |
| Comorbidity count, median [IQR] | 1 (0, 2) | 1 (0, 1) | 1 (0, 2) | 0.68 |

COVID-19, coronavirus disease 2019; SARD, systemic autoimmune rheumatic disease; csDMARD, conventional synthetic disease-modifying antirheumatic drug
\*Other conventional synthetic DMARD includes leflunomide, azathioprine, sulfasalazine, apremilast, cyclosporine, and tacrolimus
<sup>†</sup>Other biologic DMARD includes IL-6 receptor inhibitor, B-cell activating factor inhibitor, IL-23 inhibitor, IL-17 inhibitor, IL-12/IL-23 inhibitor, IL-1 inhibitor, and CTLA-4 immunoglobulin
<sup>^</sup>For patients who reported multiple SARD diagnoses, those who reported inflammatory arthritis in addition to either lupus or myositis were classified as having a “connective tissue disease (CTD)” given that inflammatory arthritis can be a component of CTD. Patients who listed both polymyalgia rheumatica and rheumatoid arthritis were classified as having “inflammatory arthritis.” Patients with multiple SARDs that can coexist (e.g., psoriatic arthritis and lupus) were classified as having multiple SARDs. In the case
of missing data or unanswered survey questions regarding rheumatic disease diagnosis or treatment, manual review of the EHR was performed to fill in this missing data.

The majority in each group were female (80% of those with breakthrough infection vs. 79% of those with non-breakthrough infection, p=0.88), and the mean age at the time of survey completion was 53 vs. 52 years, respectively (p=0.68). Most patients in each group (breakthrough versus non-breakthrough, respectively) were White (87% vs. 79%, p=0.08) and never smokers (75% vs. 71%, p=0.65). Common comorbidities in each group (breakthrough versus non-breakthrough, respectively) were also similar with the exception of obesity which was more common in those with non-breakthrough infection (25% vs. 15%, p=0.04). The median (IQR) comorbidity count was 1 (0, 1) in those with breakthrough infection compared with 1 (0, 2) in those with non-breakthrough infection (p=0.68).

The most common SARD category (in those with breakthrough versus non-breakthrough infection, respectively) was inflammatory arthritis (53% vs. 63%), followed by connective tissue disease (26% vs. 23%), vasculitis (11% vs. 8%), other disease (6% vs. 3%), or multiple diseases (4% vs. 2%) (p=0.33 for difference across categories). The most common conventional synthetic DMARDs used at the time of COVID-19 included hydroxychloroquine (29% vs. 18%, p=0.04), methotrexate (23% vs. 20%, p=0.567) and mycophenolate (12% vs. 4%, p=0.02). The most common biologic and targeted synthetic DMARDs at the time of COVID-19 infection included TNF inhibitors (24% vs. 21%, p=0.66) followed by anti-CD20 monoclonal antibodies (12% vs. 5%, p=0.08) and Janus kinase inhibitors (6% vs. 4%, p=0.40).

### Acute COVID-19 symptoms and clinical course according to breakthrough infection status

Infection during the period in which the Omicron variants were predominant (December 17, 2021 onward) was more common in patients with breakthrough COVID-19 infection (84, 72%) than in patients with non-breakthrough COVID-19 infection (3, 2%) (**Table 2**). Those with breakthrough infection had more nasal congestion/rhinorrhea (73% vs. 46%, p<0.0001) and sore throat (54% vs. 37%, p=0.01), and those with non-breakthrough infection had more anosmia (46% vs. 22%, p<0.0001), dysgeusia (45% vs. 28%, p<0.01), and joint pain (11% vs. 4%, p=0.05). Those with breakthrough infection more often received nirmatrelvir/ritonavir (12% vs. 1%, p<0.0001) and monoclonal antibody treatment (34% vs. 8%, p<0.0001) compared with those with non-breakthrough infection. Fewer patients with breakthrough COVID-19 infection required hospitalization than those with non-breakthrough infection (5% vs. 27%, p=0.001).

**Table 2.** Symptoms and clinical course for study participants with rheumatic disease and COVID-19, stratified by vaccination status at the time of infection

|  | **All rheumatic disease patients (N=280)** | **Breakthrough COVID-19 infection (N=116)** | **Non-breakthrough COVID-19 infection (N=164)** | **p-value** |
| --- | --- | --- | --- | --- |
| Date of COVID-19 diagnosis, n (%) |  |  |  | <0.0001 |
| March 1-June 30, 2020 | 45 (16) | 0 (0) | 45 (27) |  |
| July 1, 2020-January 31, 2021 | 97 (35) | 0 (0) | 97 (59) |  |
| February 1-June 30, 2021 | 21 (8) | 3 (3) | 18 (11) |  |
| July 1-December 16, 2021 | 30 (11) | 29 (25) | 1 (1) |  |
| December 17, 2021-July 8, 2022 | 87 (31) | 84 (72) | 3 (2) |  |
| COVID-19 symptoms, n (%) |  |  |  |  |
| Fatigue/malaise | 220 (79) | 93 (80) | 127 (77) | 0.66 |
| Fever | 165 (59) | 66 (57) | 99 (60) | 0.62 |
| Headache | 175 (63) | 71 (61) | 104 (63) | 0.71 |
| Myalgias | 162 (58) | 63 (54) | 99 (60) | 0.33 |
| Nasal congestion or rhinorrhea | 160 (57) | 85 (73) | 75 (46) | <0.0001 |
| Cough | 146 (52) | 73 (63) | 73 (45) | <0.01 |
| Anosmia | 101 (36) | 25 (22) | 76 (46) | <0.0001 |
| Dysgeusia | 106 (38) | 32 (28) | 74 (45) | <0.01 |
| Sore throat | 124 (44) | 63 (54) | 61 (37) | <0.01 |
| Dyspnea | 83 (30) | 28 (24) | 55 (34) | 0.11 |
| Chest pain | 59 (21) | 25 (22) | 34 (21) | 0.88 |
| Nausea or vomiting | 44 (16) | 16 (14) | 28 (17) | 0.51 |
| Joint pain | 23 (8) | 5 (4) | 18 (11) | 0.049 |
| New rash, hives, or blisters | 17 (6) | 4 (3) | 13 (8) | 0.14 |
| None | 64 (23) | 39 (34) | 25 (15) | <0.01 |
| Acute COVID-19 treatment, n (%) |  |  |  |  |
| Dexamethasone | 17 (6) | 3 (3) | 14 (9) | 0.04 |
| Remdesivir | 12 (4) | 3 (3) | 9 (5) | 0.37 |
| Monoclonal antibody | 52 (19) | 39 (34) | 13 (8) | <0.0001 |
| Nirmatrelvir/ritonavir | 15 (5) | 14 (12) | 1 (1) | <0.0001 |
| COVID-19 severity, n (%) |  |  |  | <0.01 |
| Not hospitalized | 244 (87) | 110 (95) | 134 (81) |  |
| Hospitalized with or without supplemental oxygen | 32 (11) | 5 (4) | 27 (16) |  |

COVID-19, coronavirus disease 2019; SARD, systemic autoimmune rheumatic disease; SARS-CoV-2, severe acute respiratory syndrome coronavirus 2

### Post-acute sequelae according to breakthrough infection status

Those with breakthrough infection were less likely to have PASC at 28 days (41% vs. 54%, p=0.04) and at 90 days (21% vs. 41%, p<0.0001) (**Table 3; Figure 1**), corresponding to a lower odds of PASC at 28 days (aOR 0.49, 95% CI: 0.29, 0.83) and 90 days (aOR 0.10, 95% CI: 0.04, 0.22).

**Table 3.** Duration of symptoms and post-acute sequelae of COVID-19 (PASC) following breakthrough versus non-breakthrough COVID-19 infection.

|  | **Breakthrough**<br>**COVID-19**<br>**infection**<br>**(N=116)** | **Non-**<br>**breakthrough**<br>**COVID-19**<br>**infection**<br>**(N=164)** | **p-value** |
| --- | --- | --- | --- |
| **CDC Definition of PASC (COVID-19 symptoms lasting at least 28 days post-COVID-19 infection), n (%)** | 48 (41) | 89 (54) | 0.04 |
| Unadjusted OR (95% CI) | 0.59 (0.37, 0.96) | Reference (1.0) | 0.03 |
| Adjusted OR (95% CI), Model 1<sup>†</sup> | 0.52 (0.31, 0.86) | Reference (1.0) | 0.01 |
| Adjusted OR (95% CI), Model 2<sup>‡</sup> | 0.49 (0.29, 0.83) |  | 0.01 |
| **WHO Definition of PASC (COVID-19 symptoms lasting at least 90 days post-COVID-19 infection), n (%)<sup>‡</sup>** | 10 (21) | 65 (41) | 0.01 |
| Unadjusted OR (95% CI) | 0.14 (0.07, 0.30) | Reference (1.0) | <0.0001 |
| Adjusted OR (95% CI), Model 1<sup>†</sup> | 0.11 (0.05, 0.24) | Reference (1.0) | <0.0001 |
| Adjusted OR (95% CI), Model 2<sup>‡</sup> | 0.10 (0.04, 0.22) | Reference (1.0) | <0.0001 |

CDC, Centers for Disease Control and Prevention; WHO, World Health Organization
\*Primary follow-up period was 204 days, given that this was the maximum follow-up period among those with breakthrough infections
<sup>†</sup>Multivariable model 1 is adjusted for age, sex, and race. Multivariable model 2 is adjusted for age, sex, race, comorbidity count, and use of any one of the following medications: anti-CD20 monoclonal antibodies, methotrexate, mycophenolate, or glucocorticoids.
<sup>‡</sup>Denominator includes those who completed a survey at least 90 days following COVID-19 diagnosis; N=47 with breakthrough infection and 159 with non-breakthrough infection

**Figure 1.**
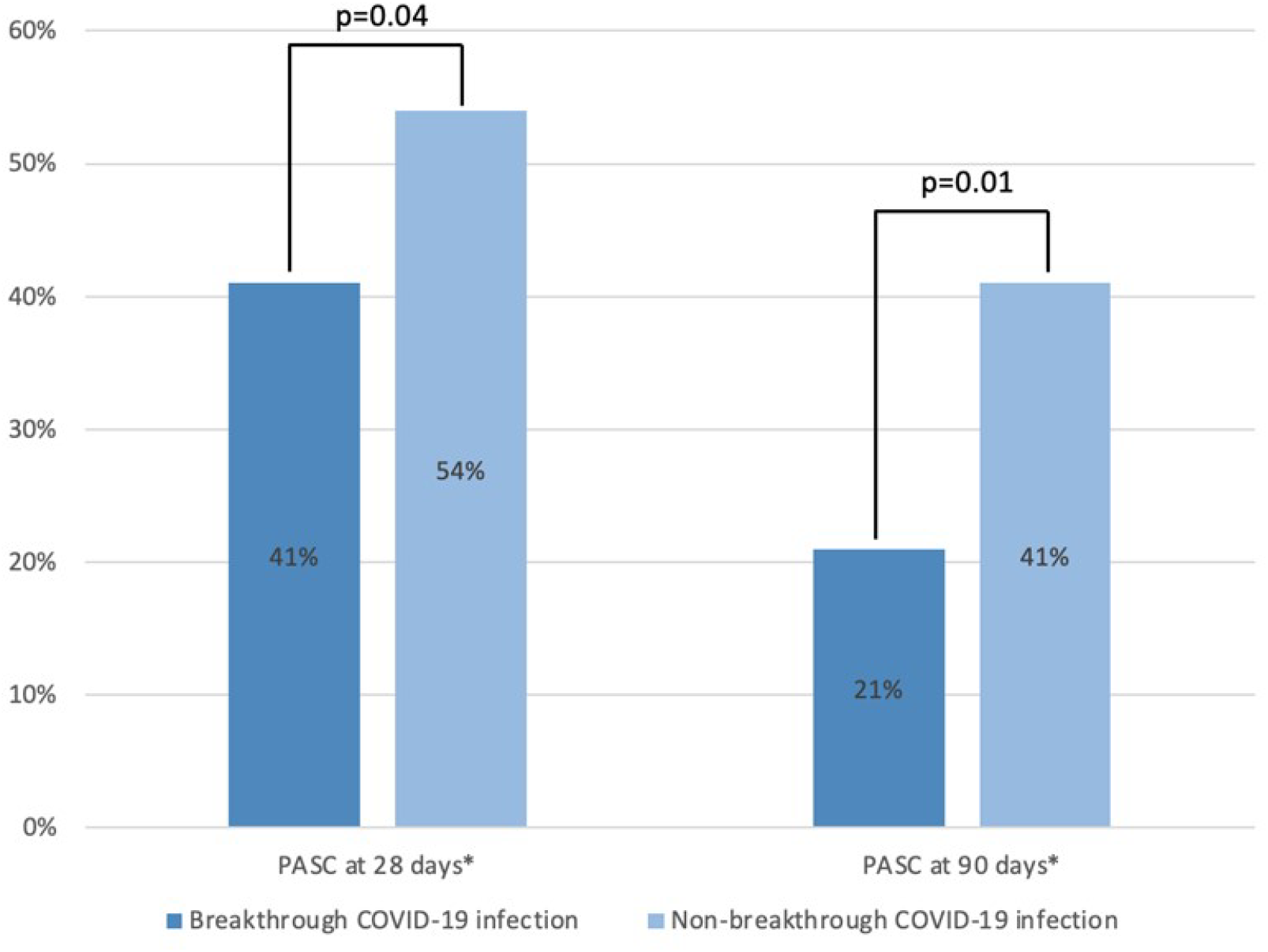
Proportion Experiencing Post-Acute Sequelae of SARS CoV-2 (PASC) among those with breakthrough and non-breakthrough COVID-19 infection. *Based on the either Centers for Disease Control and Prevention Definition of PASC (COVID-19 symptoms lasting at least 28 days post-COVID-19 infection) or the World Health Organization Definition of PASC (COVID-19 symptoms lasting at least 90 days post-COVID-19 infection) For the analysis at 90 days, the denominator includes those who completed a survey at least 90 days following COVID-19 diagnosis; N=206 overall, 47 with breakthrough infection, and 159 with non-breakthrough infection

Over 204 days, the mean time spent free from symptoms (time post-symptom resolution), reflected by the area under the cumulative incidence curves, was 137.9 days and 109.0 days, respectively, in the breakthrough and non-breakthrough groups in adjusted analyses (**Figure 2**; **Supplementary Figure 1**). Thus, the breakthrough group was symptom-free for an additional 28.9 days (95% CI: 8.83, 48.89, p=0.005) compared to the non-breakthrough group in adjusted analyses. The breakthrough group also experienced more symptom-free days over follow-up time when limited to 28 and 90 days (**Supplementary Figures 2 and 3**).

**Figure 2.**
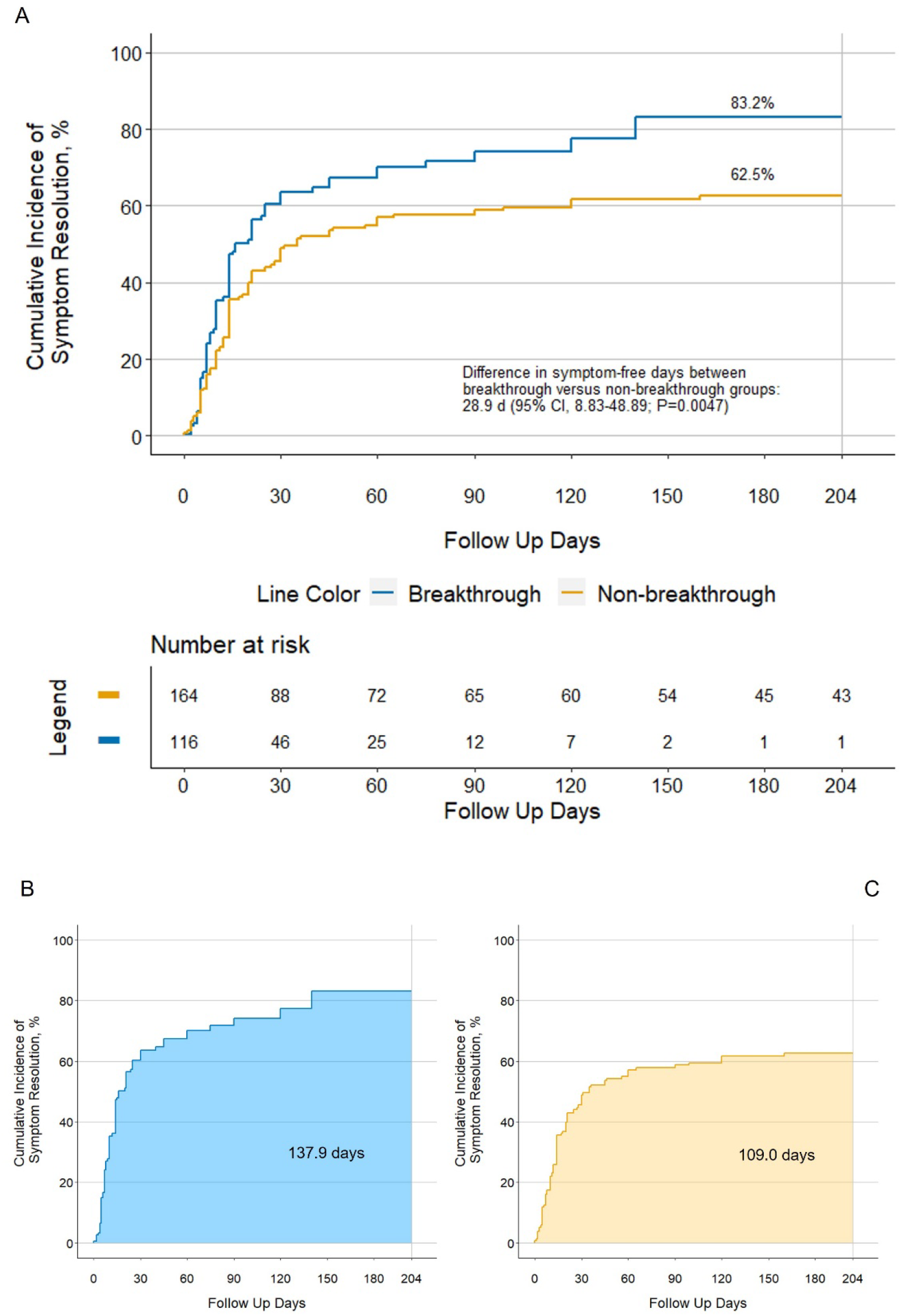
Days to symptom resolution in those with breakthrough versus non-breakthrough COVID-19 infection over 204-day follow-up period A. Cumulative incidence curves for time to symptom resolution, comparing breakthrough versus non-breakthrough infection in multivariable-adjusted analyses. B and C, mean post-symptom resolution time spans as the area under the cumulative incidence curves in those with breakthrough versus non-breakthrough infection, respectively, across 204 days of follow-up in multivariable-adjusted analyses.

Our findings remained consistent in sensitivity analyses limiting the sample to those who did not receive nirmatrelvir/ritonavir or monoclonal antibodies, those who did not receive any COVID-19-related treatment, those who completed the questionnaires within 6 months of COVID-19 infection, and those who did not require hospitalization (**Supplementary Tables 1-4**).

### Patient-reported outcomes including pain, fatigue, functional status, and rheumatic disease activity following COVID-19 infection

Pain and fatigue were less severe in those with breakthrough infection than in those with non-breakthrough infection (SF-MPQ: median score of 4 vs. 5, p=0.04 and FSI: 48 vs. 55, p=0.08, respectively) (**Figure 3A, 3B; Table 4**). Functional status (mHAQ) scores were similar between those with and without breakthrough COVID-19 infection (median of 0.1 in each group, p=0.88) (**Figure 3C**). Health-related quality of life, as assessed by the SF-12, was similar among those with and without breakthrough infection (**Figure 3D**). The median (IQR) PCS-12 was 43.6 (33.7, 52.6) in those with breakthrough infection compared with 41.0 (32.2, 49.5) in those with non-breakthrough infection (p=0.11), and the median (IQR) MCS-12 was 49.4 (41.4, 55.6) in those with breakthrough infection compared with 50.2 (37.9, 57.0) in those with non-breakthrough infection (p=0.86).

**Figure 3.**
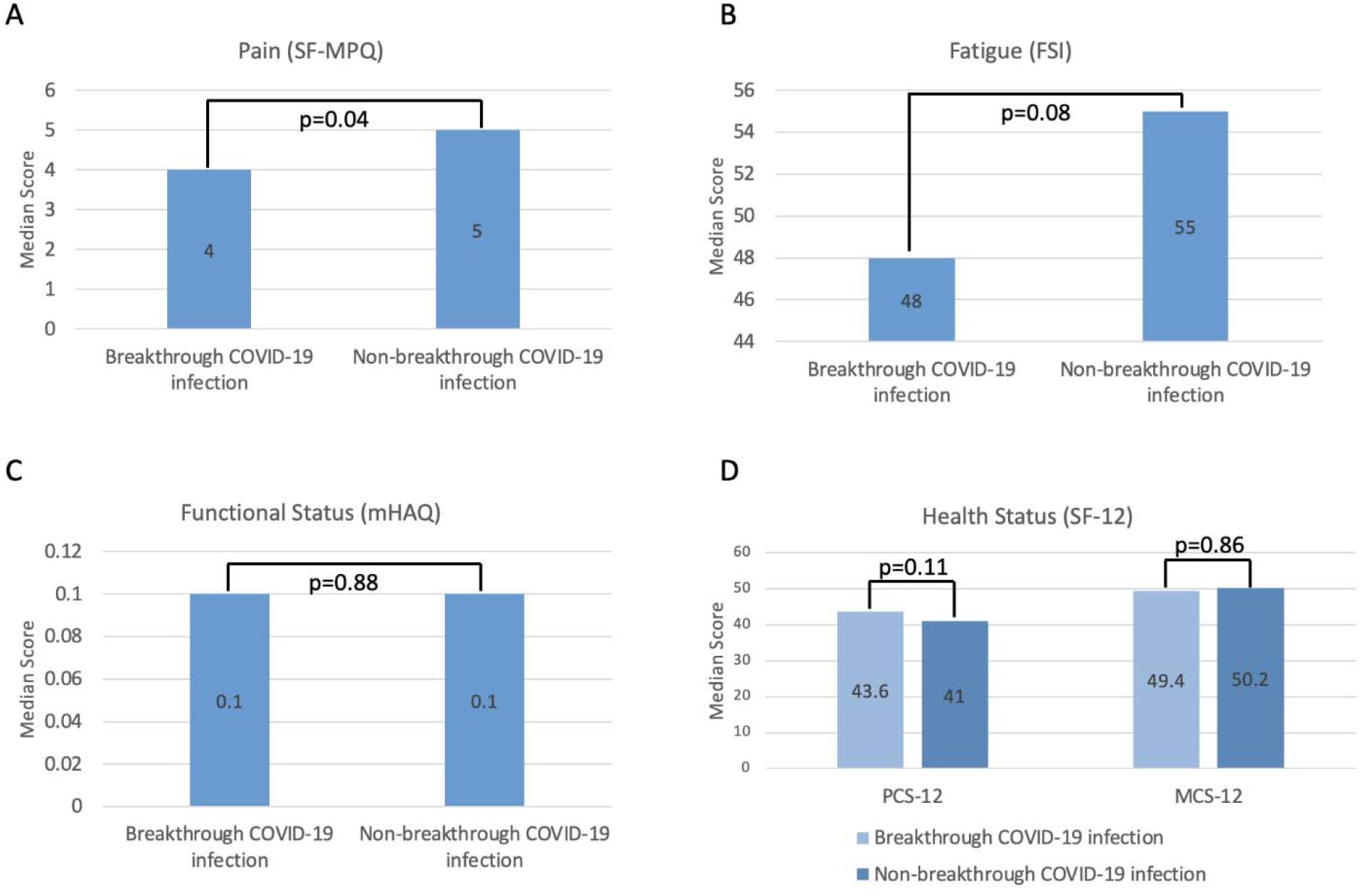
Patient-reported outcomes among those with breakthrough and non-breakthrough COVID-19 infection SF-MPQ, Short-form McGill Pain Questionnaire; FSI, Fatigue Symptom Inventory; mHAQ, modified Health Assessment Questionnaire; SF-12, Short-form 12-item survey; PCS-12, Physical Component Summary Score; PCS-12, Mental Component Summary Score. Higher SF-MPQ, FSI, and mHAQ scores indicate worse pain, fatigue, and functional status, respectively. Higher SF-12 scores, both PCS-12 and MCS-12, represent better health status.

**Table 4.** Patient-reported outcomes in rheumatic disease patients after COVID-19 infection, based on self-report at the time of survey completion.

|  | **All**<br>**rheumatic**<br>**disease**<br>**patients**<br>**(N=280)** | **Breakthrough**<br>**COVID-19**<br>**infection**<br>**(N=116)** | **Non-**<br>**breakthrough**<br>**COVID-19**<br>**infection**<br>**(N=164)** | **p-value** |
| --- | --- | --- | --- | --- |
| Pain (SF-MPQ) |  |  |  |  |
| N | 258 | 111 | 147 |  |
| Median score [IQR] | 4 (2, 10) | 4 (1, 8) | 5 (3, 10) | 0.04 |
| Pain rating index ordinal<br>category, n (%) |  |  |  | 0.20 |
| No pain | 41 (15) | 17 (15) | 24 (15) |  |
| Mild pain | 86 (31) | 43 (37) | 43 (26) |  |
| Discomforting, distressing,<br>horrible, or excruciating pain | 135 (48) | 51 (44) | 84 (51) |  |
| Fatigue (FSI) |  |  |  |  |
| N | 264 | 111 | 153 |  |
| Median score [IQR] | 54 (26, 82) | 48 (21, 79) | 55 (28, 84) | 0.08 |
| Functional status (mHAQ) |  |  |  |  |
| N | 270 | 112 | 158 |  |
| Median score [IQR] | 0.1 (0.0, 0.5) | 0.1 (0.0, 0.5) | 0.1 (0.0, 0.5) | 0.88 |
| Categorical score, n (%) |  |  |  | 0.61 |
| Normal (<0.3) | 163 (58) | 70 (60) | 93 (57) |  |
| Mild, moderate, or severe (0.3<br>to >1.8) | 107 (38) | 42 (36) | 65 (40) |  |
| Health status (SF-12) |  |  |  |  |
| PCS-12 score, median [IQR] | 42.1 (33.0,<br>50.8) | 43.6 (33.7, 52.6) | 41.0 (32.2, 49.5) | 0.11 |
| MCS-12 score, median [IQR] | 46.8 (36.9,<br>54.4) | 49.3 (41.4, 55.6) | 50.2 (37.9, 57.0) | 0.86 |

CDC, Centers for Disease Control and Prevention; WHO, World Health Organization; FSI, Fatigue Symptom Inventory; mHAQ, modified Health Assessment Questionnaire; SF-12, Short-form 12-item survey; SF-MPQ, Short-form McGill Pain Questionnaire; PCS-12, Physical Component Summary score; MCS-12, Mental Component Summary score

Patient-reported outcome measures comparing PASC following breakthrough COVID-19 infection (n=48) versus PASC following non-breakthrough COVID-19 infection (n=89) were similar in terms of pain, fatigue, functional status, and overall health status (**Supplementary Table 5**). The frequency and timing relative to infection of self-reported flares of the underlying SARD were also similar following COVID-19 infection in those with breakthrough versus non-breakthrough infection (40% vs. 42%, p=0.71) (**Supplementary Table 6**).

## Discussion

In this prospective study of patients with SARDs and COVID-19, those with breakthrough infection had significantly shorter symptom duration and lower rates of PASC than those unvaccinated or partially vaccinated prior to infection. This corresponded with less pain and fatigue, two common manifestations of PASC, in those with breakthrough infection compared to those with non-breakthrough infection following the acute course. Collectively, our findings suggest that SARS-CoV-2 vaccination reduces the risk of PASC in SARD patients, in addition to the known reduction in the risk of severe acute COVID-19 outcomes. These results provide further rationale for vaccination among SARD patients.

There are limited data regarding the potential impact of SARS-CoV-2 vaccination on the risk of PASC in the general population and, to our knowledge, no studies in patients with SARDs. A previous community-based study of the general population in the United Kingdom found a nearly 50% reduced risk of PASC (≥ 28 days) in those with a breakthrough infection (OR 0.51, 95% CI: 0.32, 0.82).^16^ Similar findings were observed in a cohort study of the Israeli general population after COVID-19.^33^ Further, a large study conducted among Veterans Affairs beneficiaries found that the risk of cardiovascular, pulmonary, metabolic, and coagulopathic sequelae was lower in those with breakthrough COVID-19.^17^ Our findings expand upon these prior studies, providing important new evidence in SARD patients suggesting that despite concerns regarding the impact of SARD diagnoses and their treatments on vaccine immunogenicity, vaccination provides important long-term benefits after acute COVID-19.

Due to the timing of introduction of SARS-CoV-2 vaccines, calendar time varied between those with and without breakthrough infection. Those with breakthrough infection were more often infected later in the pandemic when the Delta and Omicron variants were predominant. It is therefore possible that our findings may the result of differences in the SARS-CoV-2 variants rather than the effects of vaccination. However, previous studies have suggested that severity of COVID-19 is not intrinsically lower with Omicron versus earlier variants.^34^ Rather, differences in severity are likely due to rates of vaccination and immunity from prior infection. Because of the high rates of early vaccination among patients with SARDs, we are unable to compare the rates of PASC among those with and without breakthrough infection during time periods characterized by the predominance of a single SARS-CoV-2 variant. Other improvements during the pandemic, such as outpatient treatment, may also impact our findings, though our findings remained consistent in sensitivity analyses excluding these patients.

Importantly, PASC remained relatively common (41% with symptoms lasting ≥28 days) among those with breakthrough infection, highlighting the ongoing need to better understand the etiology of PASC in SARD patients and to identify effective treatments for PASC. Indeed, the severity of PASC, as measured by validated assessments of fatigue, pain, disability, and health-related quality of life of those with PASC, was similar regardless of whether it was associated with a breakthrough or a non-breakthrough infection. Therefore, once a SARD patient develops PASC, the severity does not appear to be influenced by vaccination status. The etiology of PASC remains unknown but several factors have been hypothesized to influence risk, including alterations in inflammatory cytokine profiles, cellular immune responses, reactivation of chronic viral infections, and autoantibody formation.^35-41^ Vaccination may reduce the risk of PASC by shortening the duration of viremia, reducing the risk of severe COVID-19 and the associated hyperinflammatory state, influencing the cellular immune response to acute infection, among other possible explanations.

Our study has several strengths. First, we used a systematic approach to identify patients with a prevalent diagnosis of a SARD at the time of SARS-CoV-2 infection. Second, we prospectively enrolled patients in RheumCARD to assess symptom duration and vaccination status, collecting patient-reported outcomes unavailable from EHR data. Third, we used two complementary definitions of PASC and conducted multiple sensitivity analyses to confirm the robustness of our findings.

Despite these strengths, our study has certain limitations. First, this study was conducted among participants who receive their care at Mass General Brigham which may limit generalizability to more diverse populations. Second, the time between COVID-19 infection and survey completion was shorter among those with breakthrough infection because of the timing of the initiation of RheumCARD. While this could introduce recall bias whereby those with a non-breakthrough infection reported a longer duration of symptoms, we do not have reason to suspect this is likely. Further, our findings were similar in a sensitivity analysis where we limited the analysis to those who completed the surveys within six months of their index date. Also, similar proportions of patients in each group recalled flares of their underlying SARD following COVID-19 infection, suggesting no significant differential recall bias according to vaccination status. Third, some patients in the breakthrough group also received antiviral and other COVID-19 treatments which could impact our findings. However, in sensitivity analyses we found that our observed trends persisted despite accounting for these differences. Fourth, based on the timing of infection, the variants most prevalent in those with breakthrough infections were Delta and Omicron. Due to the high vaccination rate in our cohort once vaccines became available, we are unable to compare those with and without breakthrough infection with the same SARS-CoV-2 variant. However, previous studies do not suggest that there are differences in the risk of PASC based on variant. Finally, while we used the currently accepted research definitions of PASC, it is possible that the patient reports could reflect underlying SARD activity, organ damage, or be otherwise unrelated to COVID-19 resulting in overestimation of people truly experiencing prolonged COVID-19 symptoms. However, these factors are unlikely to be differentially related to vaccine status and are unlikely to explain our findings. Future studies are needed to determine whether more homogeneous PASC subtypes may be present and to elucidate pathogenesis and possible treatments.

In conclusion, we found that patients with SARDs have shorter duration of COVID-19 symptoms and are less likely to have PASC at both 28 and 90 days if they are fully vaccinated prior to acute infection. These findings suggest that despite a higher risk of breakthrough infection, vaccination in patients with SARDs not only reduces the risk of severe acute outcomes but also long-term outcomes. Nonetheless, PASC remains common among SARD patients, even after vaccination, and when present, the severity is similar to those who were either unvaccinated or partially vaccinated. Additional investigation is needed to determine the etiology and effective treatments of PASC in SARDs.

## Data Availability

All data produced in the present study are available upon reasonable request to the authors

