## Supplemental Material for "Impact of vaccination on post-acute sequelae of SARS CoV-2 infection in patients with rheumatic diseases"

**Supplementary Table 1**. Persistent COVID-19 symptoms and other patient-reported outcomes in rheumatic disease patients following COVID-19 infection among those who completed surveys within 6 months of COVID-19 infection.

|  | **All rheumatic disease patients (N=147)** | **Breakthrough COVID-19 infection (N=106)** | **Non-breakthrough COVID-19 infection (N=41)** | **p-value** |
| --- | --- | --- | --- | --- |
| Days to acute symptom resolution or date of survey if symptoms ongoing, median [IQR] | 21 (8, 60) | 21 (8, 56) | 25 (10, 91) | 0.23 |
| CDC Definition of PASC (COVID-19 symptoms lasting at least 28 days post-COVID-19 infection) | 66 (45) | 46 (43) | 20 (49) | 0.58 |
| WHO Definition of PASC (COVID-19 symptoms lasting at least 90 days post-COVID-19 infection)* | 21 (29) | 9 (24) | 12 (33) | 0.40 |
| Pain (SF-MPQ) |  |  |  |  |
| N | 139 | 101 | 38 |  |
| Median score [IQR] | 4 (1, 9) | 4 (1, 9) | 4.5 (2, 9) | 0.62 |
| Pain rating index ordinal categories, n (%) |  |  |  | 0.20 |
| No pain | 24 (16) | 14 (13) | 10 (24) |  |
| Mild pain | 47 (32) | 37 (35) | 10 (24) |  |
| Discomforting, distressing, horrible, or excruciating pain | 69 (47) | 50 (47) | 19 (46) |  |
| Fatigue (FSI) |  |  |  |  |
| N | 140 | 101 | 39 |  |
| Median score [IQR] | 54 (25, 82.5) | 51 (23, 81) | 58 (27, 85) | 0.35 |
| Functional status (mHAQ) |  |  |  |  |
| N | 142 | 103 | 39 |  |
| Median score [IQR] | 0.1 (0, 0.5) | 0.1 (0, 0.5) | 0.1 (0, 0.4) | 0.51 |
| Functional status categorical score, n (%) |  |  |  | 1.00 |
| Normal (<0.3) | 89 (61) | 64 (60) | 25 (61) |  |
| Mild, moderate, or severe (0.3 to >1.8) | 53 (36) | 39 (37) | 14 (34) |  |
| Health status (SF-12) |  |  |  |  |
| PCS-12 score, median [IQR] | 42.3 (33.4, 51.1) | 43.6 (33.4, 52.5) | 40.8 (33.4, 47.5) | 0.43 |
| MCS-12 score, median [IQR] | 49.2 (39.4, 55.5) | 49 (41.3, 55) | 49.9 (35.8, 57.6) | 0.79 |

CDC, Centers for Disease Control and Prevention; WHO, World Health Organization; FSI, Fatigue Symptom Inventory; mHAQ, modified Health Assessment Questionnaire; SF-12, Short-form 12-item survey; SF-MPQ, Short-form McGill Pain Questionnaire; PCS-12, Physical Component Summary score; MCS-12, Mental Component Summary score

*Denominator includes those who completed a survey at least 90 days following COVID-19 diagnosis; N=73 overall, 37 with breakthrough infection, and 36 with non-breakthrough infection

**Supplementary Table 2**. Persistent COVID-19 symptoms and other patient-reported outcomes in rheumatic disease patients following COVID-19 infection among those who did not receive either monoclonal antibodies or nirmatrelvir/ritonavir

|  | **All rheumatic disease patients (N=214)** | **Breakthrough COVID-19 infection (N=64)** | **Non-breakthrough COVID-19 infection (N=150)** | **p-value** |
| --- | --- | --- | --- | --- |
| Days to acute symptom resolution or date of survey if symptoms ongoing, median [IQR] | 30 (10, 120) | 25 (8, 57) | 30 (12, 216) | 0.01 |
| CDC Definition of PASC (COVID-19 symptoms lasting at least 28 days post-COVID-19 infection) | 108 (50) | 27 (42) | 81 (54) | 0.14 |
| WHO Definition of PASC (COVID-19 symptoms lasting at least 90 days post-COVID-19 infection)* | 60 (35) | 3 (9) | 57 (51) | 0.01 |
| Pain (SF-MPQ) |  |  |  |  |
| N | 193 | 60 | 133 |  |
| Median score [IQR] | 5 (2, 10) | 3 (1, 7) | 6 (3, 10) | 0.02 |
| Pain rating index ordinal categories, n (%) |  |  |  | 0.05 |
| No pain | 31 (14) | 8 (13) | 23 (15) |  |
| Mild pain | 64 (30) | 27 (42) | 37 (25) |  |
| Discomforting, distressing, horrible, or excruciating pain | 102 (48) | 25 (39) | 77 (51) |  |
| Fatigue (FSI) |  |  |  |  |
| N | 199 | 60 | 139 |  |
| Median score [IQR] | 51 (25, 80) | 36 (14, 68) | 55 (28, 84) | 0.02 |
| Functional status (mHAQ) |  |  |  |  |
| N | 205 | 61 | 144 |  |
| Median score [IQR] | 0.1 (0.0, 0.4) | 0.1 (0.0, 0.4) | 0.2 (0.0, 0.5) | 0.42 |
| Functional status categorical score, n (%) |  |  |  | 0.08 |
| Normal (<0.3) | 128 (60) | 44 (69) | 84 (56) |  |
| Mild, moderate, or severe (0.3 to >1.8) | 77 (36) | 17 (27) | 60 (40) |  |
| Health status (SF-12) |  |  |  |  |
| PCS-12 score, median [IQR] | 42.3 (32.8, 51.0) | 44.6 (36.4, 53.9) | 41.1 (32.1, 49.9) | 0.09 |
| MCS-12 score, median [IQR] | 49.9 (39.1, 56.3) | 49.1 (39.7, 56.0) | 50.7 (37.8, 57.1) | 0.98 |

CDC, Centers for Disease Control and Prevention; WHO, World Health Organization; FSI, Fatigue Symptom Inventory; mHAQ, modified Health Assessment Questionnaire; SF-12, Short-form 12-item survey; SF-MPQ, Short-form McGill Pain Questionnaire; PCS-12, Physical Component Summary score; MCS-12, Mental Component Summary score

*Denominator includes those who completed a survey at least 90 days following COVID-19 diagnosis; N=171 overall, 26 with breakthrough infection, and 145 with non-breakthrough infection

**Supplementary Table 3**. Persistent COVID-19 symptoms and other patient-reported outcomes in rheumatic disease patients following COVID-19 infection among those who did not receive any COVID-19-specific therapy.

|  | **All rheumatic disease patients (N=192)** | **Breakthrough COVID-19 infection (N=60)** | **Non-breakthrough COVID-19 infection (N=132)** | **p-value** |
| --- | --- | --- | --- | --- |
| Days to acute symptom resolution or date of survey if symptoms ongoing, median [IQR] | 25 (10, 92) | 23 (8, 57) | 28 (12, 200) | 0.02 |
| CDC Definition of PASC (COVID-19 symptoms lasting at least 28 days post-COVID-19 infection) | 90 (47) | 24 (40) | 66 (50) | 0.22 |
| WHO Definition of PASC (COVID-19 symptoms lasting at least 90 days post-COVID-19 infection)* | 48 (32) | 3 (8) | 45 (40) | 0.03 |
| Pain (SF-MPQ) |  |  |  |  |
| N | 174 | 57 | 117 |  |
| Median score [IQR] | 5 (2, 10) | 3 (1, 7) | 6 (3, 11) | 0.02 |
| Pain rating index ordinal categories, n (%) |  |  |  | 0.02 |
| No pain | 27 (14) | 7 (12) | 20 (15) |  |
| Mild pain | 58 (30) | 27 (45) | 31 (23) |  |
| Discomforting, distressing, horrible, or excruciating pain | 93 (48) | 23 (38) | 70 (53) |  |
| Fatigue (FSI) |  |  |  |  |
| N | 180 | 57 | 123 |  |
| Median score [IQR] | 49 (24, 80) | 33 (13, 66) | 54 (27, 84) | 0.04 |
| Functional status (mHAQ) |  |  |  |  |
| N | 184 | 57 | 127 |  |
| Median score [IQR] | 0.1 (0.0, 0.4) | 0.1 (0.0, 0.4) | 0.1 (0.0, 0.5) | 0.25 |
| Functional status categorical score, n (%) |  |  |  | 0.07 |
| Normal (<0.3) | 117 (61) | 42 (70) | 75 (57) |  |
| Mild, moderate, or severe (0.3 to >1.8) | 67 (35) | 15 (25) | 52 (39) |  |
| Health status (SF-12) |  |  |  |  |
| PCS-12 score, median [IQR] | 42.7 (33.2, 51.7) | 45.5 (36.4, 54.0) | 41.1 (32.6, 50.3) | 0.14 |
| MCS-12 score, median [IQR] | 50.9 (39.3, 57.0) | 49.4 (39.6, 56.0) | 51.3 (38.0, 57.6) | 0.90 |

CDC, Centers for Disease Control and Prevention; WHO, World Health Organization; FSI, Fatigue Symptom Inventory; mHAQ, modified Health Assessment Questionnaire; SF-12, Short-form 12-item survey; SF-MPQ, Short-form McGill Pain Questionnaire; PCS-12, Physical Component Summary score; MCS-12, Mental Component Summary score

*Denominator includes those who completed a survey at least 90 days following COVID-19 diagnosis; N=152 overall, 24 with breakthrough infection, and 128 with non-breakthrough infection

**Supplementary Table 4**. Persistent COVID-19 symptoms and other patient-reported outcomes in rheumatic disease patients following COVID-19 infection among those who did not require hospitalization for COVID-19 infection.

|  | **All rheumatic disease patients (N=244)** | **Breakthrough COVID-19 infection (N=110)** | **Non-breakthrough COVID-19 infection (N=134)** | **p-value** |
| --- | --- | --- | --- | --- |
| Days to acute symptom resolution or date of survey if symptoms ongoing, median [IQR] | 21 (10, 84) | 21 (8, 51) | 25 (10, 176) | 0.01 |
| CDC Definition of PASC (COVID-19 symptoms lasting at least 28 days post-COVID-19 infection) | 112 (46) | 46 (42) | 66 (49) | 0.25 |
| WHO Definition of PASC (COVID-19 symptoms lasting at least 90 days post-COVID-19 infection)* | 57 (33) | 10 (23) | 47 (36) | 0.12 |
| Pain (SF-MPQ) |  |  |  |  |
| N | 224 | 106 | 118 |  |
| Median score [IQR] | 4 (2, 10) | 4 (1, 8) | 5 (2, 10) | 0.15 |
| Pain rating index ordinal categories, n (%) |  |  |  | 0.53 |
| No pain | 35 (14) | 16 (15) | 19 (14) |  |
| Mild pain | 77 (32) | 41 (37) | 36 (27) |  |
| Discomforting, distressing, horrible, or excruciating pain | 116 (48) | 49 (45) | 67 (50) |  |
| Fatigue (FSI) |  |  |  |  |
| N | 230 | 106 | 124 |  |
| Median score [IQR] | 52 (25, 81) | 49 (21, 79) | 54 (28, 84) | 0.18 |
| Functional status (mHAQ) |  |  |  |  |
| N | 235 | 106 | 129 |  |
| Median score [IQR] | 0.1 (0.0, 0.5) | 0.1 (0.0, 0.5) | 0.1 (0.0, 0.4) | 0.62 |
| Functional status categorical score, n (%) |  |  |  | 0.87 |
| Normal (<0.3) | 145 (59) | 66 (60) | 79 (59) |  |
| Mild, moderate, or severe (0.3 to >1.8) | 90 (37) | 40 (36) | 50 (37) |  |
| Health status (SF-12) |  |  |  |  |
| PCS-12 score, median [IQR] | 42.8 (33.6, 51.7) | 43.6 (33.7, 53.5) | 42.2 (33.4, 50.3) | 0.48 |
| MCS-12 score, median [IQR] | 49.9 (39.3, 56.0) | 49.0 (41.3, 55.4) | 51.2 (36.7, 57.7) | 0.98 |

CDC, Centers for Disease Control and Prevention; WHO, World Health Organization; FSI, Fatigue Symptom Inventory; mHAQ, modified Health Assessment Questionnaire; SF-12, Short-form 12-item survey; SF-MPQ, Short-form McGill Pain Questionnaire

*Denominator includes those who had completed a survey at least 90 days following COVID-19 diagnosis; N=172 overall, 43 with breakthrough infection, and 129 with non-breakthrough infection

**Supplementary Table 5.** Patient-reported outcomes among patients with PASC following breakthrough versus non-breakthrough COVID-19 infection

| **Characteristic** | **All patients with PASC (N=137)** | **PASC following breakthrough COVID-19 infection (N=48)** | **PASC following non-breakthrough COVID-19 infection (N=89)** | **p-value** |
| --- | --- | --- | --- | --- |
| Pain (SF-MPQ) |  |  |  |  |
| N | 124 | 45 | 79 |  |
| Median score [IQR] | 7 (3, 12) | 6 (2, 13) | 7 (3, 11) | 0.74 |
| Pain rating index ordinal categories, n (%) |  |  |  | 0.32 |
| No pain | 12 (9) | 2 (4) | 10 (11) |  |
| Mild pain | 31 (23) | 13 (27) | 18 (20) |  |
| Discomforting, distressing, horrible, or excruciating pain | 83 (61) | 30 (63) | 53 (60) |  |
| Fatigue (FSI) |  |  |  |  |
| N | 128 | 45 | 83 |  |
| Median score [IQR] | 67 (36, 90.5) | 66 (33, 89) | 67 (36, 92) | 0.66 |
| Functional status (mHAQ) |  |  |  |  |
| N | 129 | 45 | 84 |  |
| Median score [IQR] | 0.4 (0.0, 0.8) | 0.3 (0.1, 0.8) | 0.4 (0.0, 0.7) | 0.32 |
| Functional status categorical score, n (%) |  |  |  | 0.85 |
| Normal (<0.3) | 64 (47) | 23 (48) | 41 (46) |  |
| Mild, moderate, or severe (0.3 to >1.8) | 65 (47) | 22 (46) | 43 (48) |  |
| Health status (SF-12) |  |  |  |  |
| Physical Component Summary (PCS-12) score, median [IQR] | 38.3 (31.4, 47.4) | 37.6 (31.5, 46.1) | 38.8 (31.2, 48.1) | 0.70 |
| Mental Component Summary (MCS-12) score, median [IQR] | 46.4 (38, 54.9) | 45.5 (39.5, 54.0) | 46.8 (36.7, 55.8) | 0.63 |
| Persistent symptoms at time of survey, n (%) |  |  |  |  |
| Fatigue/malaise | 39 (28) | 16 (33) | 23 (26) | 0.43 |
| Anosmia | 25 (18) | 6 (13) | 19 (21) | 0.25 |
| Dysgeusia | 22 (16) | 5 (10) | 17 (19) | 0.23 |
| Dyspnea and/or cough | 15 (11) | 4 (8) | 11 (12) | 0.57 |
| Nasal congestion or rhinorrhea | 17 (12) | 8 (17) | 9 (10) | 0.29 |
| Headache | 20 (15) | 11 (23) | 9 (10) | 0.07 |
| Myalgias | 16 (12) | 8 (17) | 8 (9) | 0.26 |
| Fever | 3 (2) | 0 (0) | 3 (3) | 0.55 |
| Sore throat | 5 (4) | 2 (4) | 3 (3) | 1.00 |

SF-MPQ, Short-form McGill Pain Questionnaire; FSI, Fatigue Symptom Inventory; mHAQ, modified Health Assessment Questionnaire; SF-12, Short-form 12-item survey

**Supplementary Table 6.** Self-reported systemic rheumatic disease activity and flare following COVID-19 infection.

|  | **All rheumatic disease patients (N=280)** | **Breakthrough COVID-19 infection (N=116)** | **Non-breakthrough COVID-19 infection (N=164)** | **p-value** |
| --- | --- | --- | --- | --- |
| Self-reported SARD flare after COVID-19, n (%) | 115 (41) | 46 (40) | 69 (42) | 0.71 |
| Timing of self-reported SARD flare after COVID-19, n (%) |  |  |  | 0.19 |
| <1 week | 32 (11) | 17 (15) | 15 (9) |  |
| 1-4 weeks | 49 (18) | 20 (17) | 29 (18) |  |
| 4-12 weeks | 26 (9) | 7 (6) | 19 (12) |  |
| >12 weeks | 8 (3) | 2 (2) | 6 (4) |  |
| Participant global assessment of disease activity before COVID-19 onset, mean ± SD | 7.6 (2.3) | 7.6 (2.4) | 7.6 (2.3) | 0.98 |
| Participant global assessment of disease activity at time of survey, mean ± SD | 6.7 (2.7) | 6.8 (2.5) | 6.6 (2.8) | 0.41 |
| Disease activity by RAPID-3 at time of survey |  |  |  |  |
| Median score [IQR] | 3.0 (1.3, 4.7) | 3.0 (1.3, 4.7) | 3.3 (1.3, 4.7) | 0.48 |
| Categorical score, n (%) |  |  |  | 0.86 |
| Remission (0) | 17 (6) | 6 (5) | 11 (7) |  |
| Near remission (0.3-1.0) | 37 (13) | 17 (15) | 20 (12) |  |
| Low severity (1.3-2.0) | 34 (12) | 13 (11) | 21 (13) |  |
| Moderate severity (2.3-4.0) | 84 (30) | 31 (27) | 53 (32) |  |
| High severity (4.3-10.0) | 82 (29) | 29 (25) | 53 (32) |  |

COVID-19, coronavirus disease 2019; RAPID3, Routine Assessment of Participant Index Data 3; SARD, systemic autoimmune rheumatic disease

**Supplementary Figure 1**. Days to symptom resolution in those with breakthrough versus non-breakthrough COVID-19 infection over 204-day follow-up period (Unadjusted Analysis)

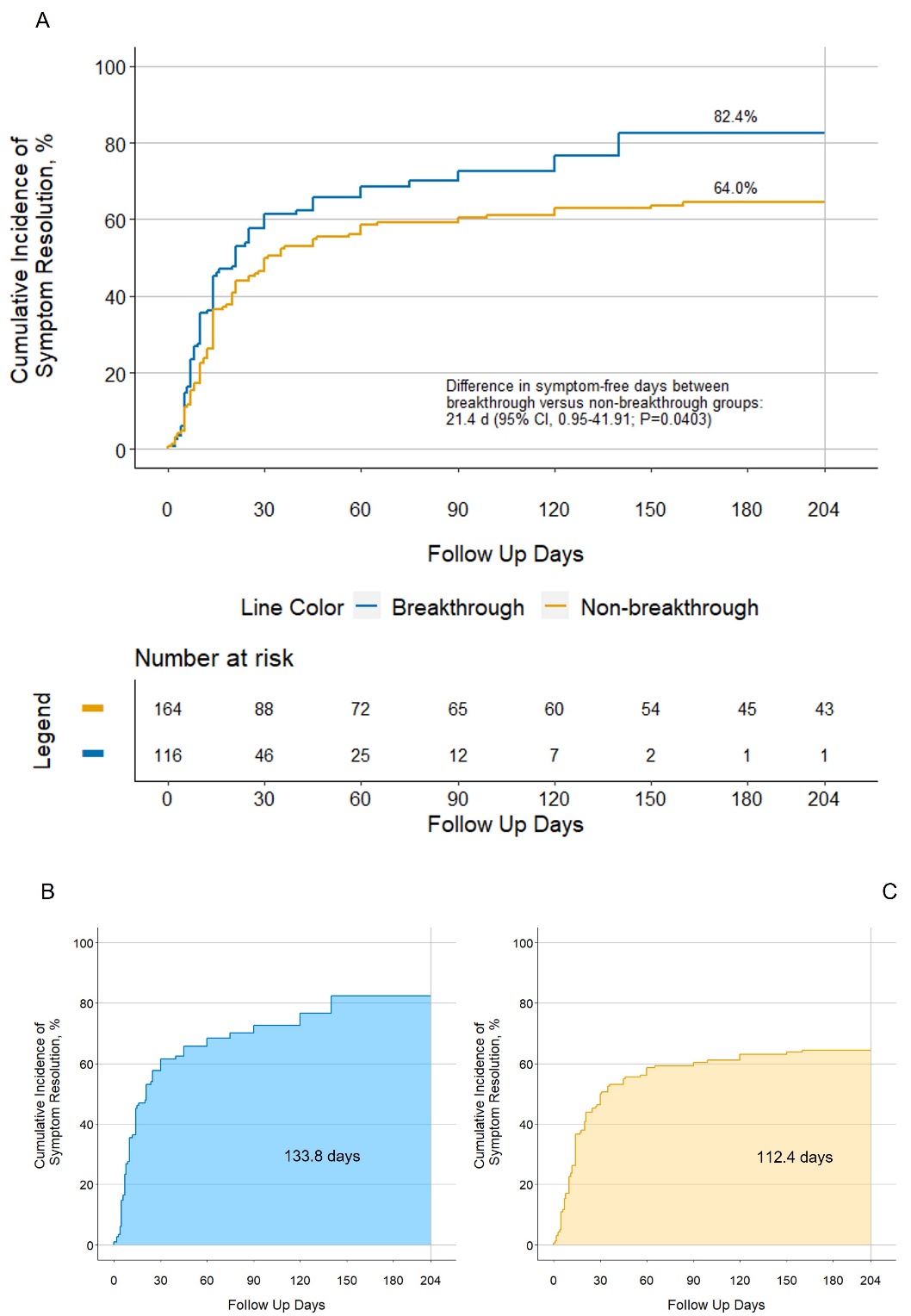

A. Cumulative incidence curves for time to symptom resolution, comparing breakthrough versus non-breakthrough infection. B and C, mean post-symptom resolution time spans as the area under the cumulative incidence curves in those with breakthrough versus non-breakthrough infection, respectively, across 204 days of follow-up.

**Supplementary Figure 2A**. Days to symptom resolution in those with breakthrough versus non-breakthrough COVID-19 infection over 28-day follow-up period (Unadjusted Analysis)

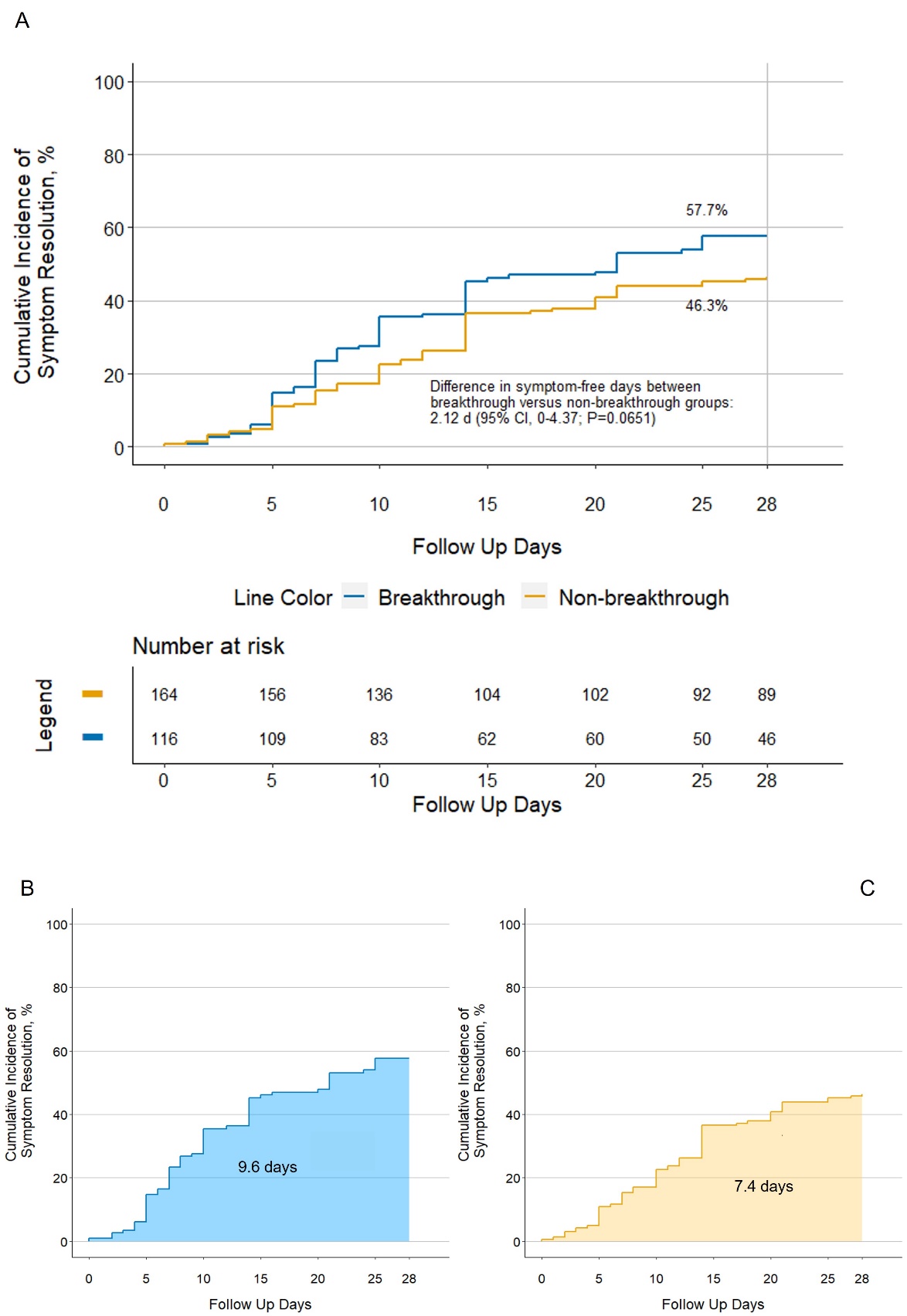

A. Cumulative incidence curves for time to symptom resolution, comparing breakthrough versus non-breakthrough infection. B and C, mean post-symptom resolution time spans as the area under the cumulative incidence curves in those with breakthrough versus non-breakthrough infection, respectively, across 28 days of follow-up.

**Supplementary Figure 2B**. Days to symptom resolution in those with breakthrough versus non-breakthrough COVID-19 infection over 28-day follow-up period (Adjusted Analysis)

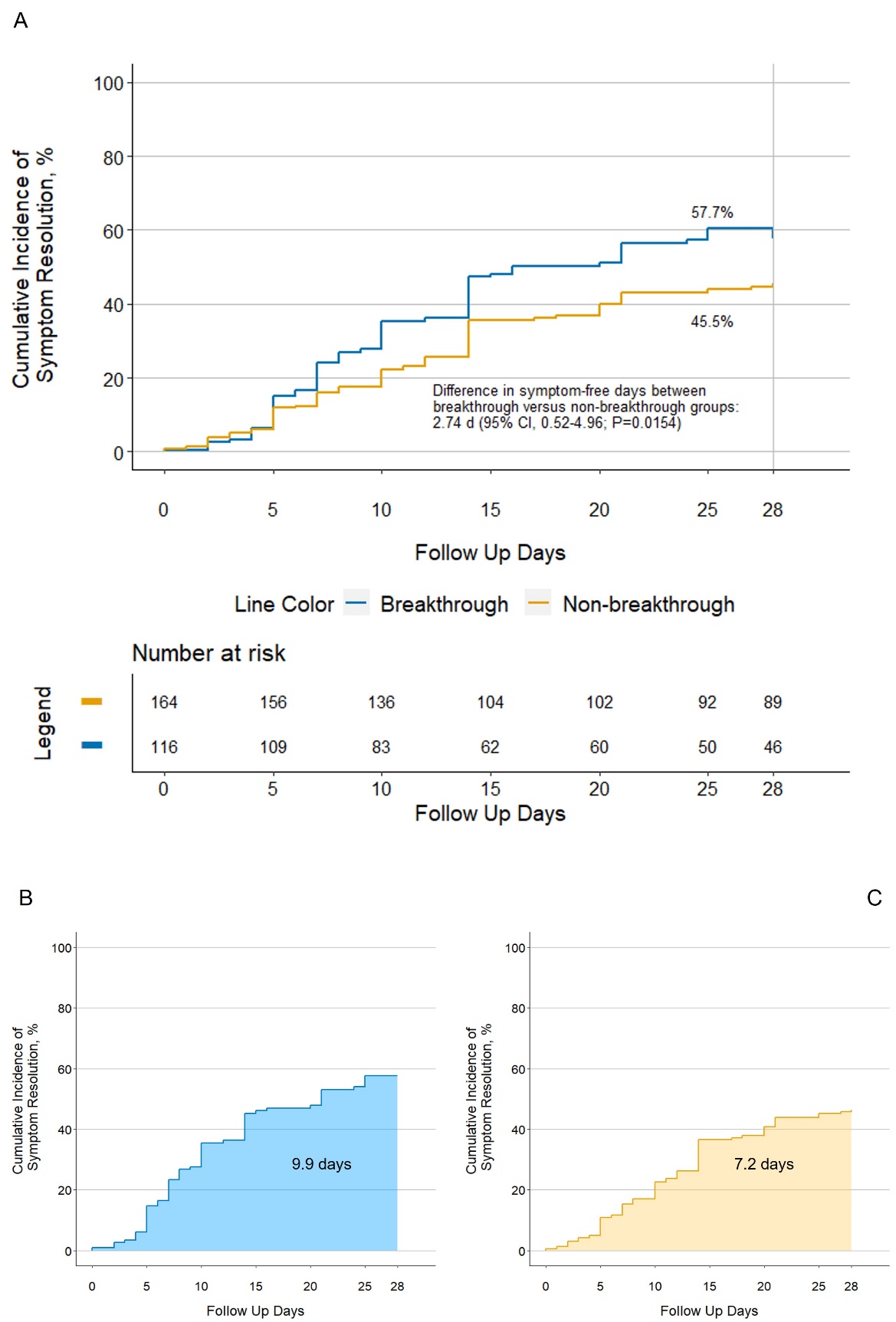

A. Cumulative incidence curves for time to symptom resolution, comparing breakthrough versus non-breakthrough infection. B and C, mean post-symptom resolution time spans as the area under the cumulative incidence curves in those with breakthrough versus non-breakthrough infection, respectively, across 28 days of follow-up.

**Supplementary Figure 3A**. Days to symptom resolution in those with breakthrough versus non-breakthrough COVID-19 infection over 90-day follow-up period (Unadjusted Analysis)

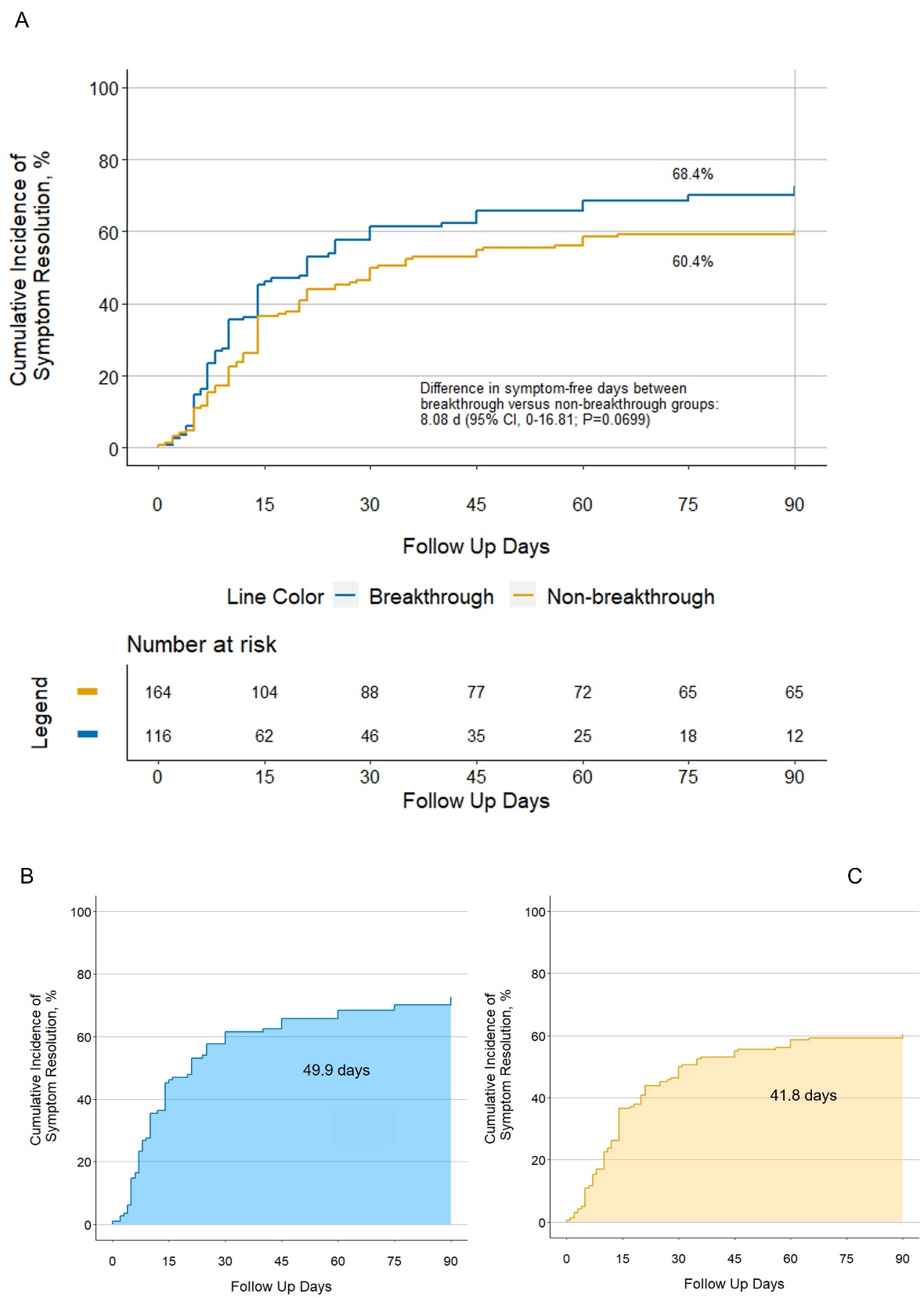

A. Cumulative incidence curves for time to symptom resolution, comparing breakthrough versus non-breakthrough infection. B and C, mean post-symptom resolution time spans as the area under the cumulative incidence curves in those with breakthrough versus non-breakthrough infection, respectively, across 90 days of follow-up.

**Supplementary Figure 3B**. Days to symptom resolution in those with breakthrough versus non-breakthrough COVID-19 infection over 90-day follow-up period (Adjusted Analysis)

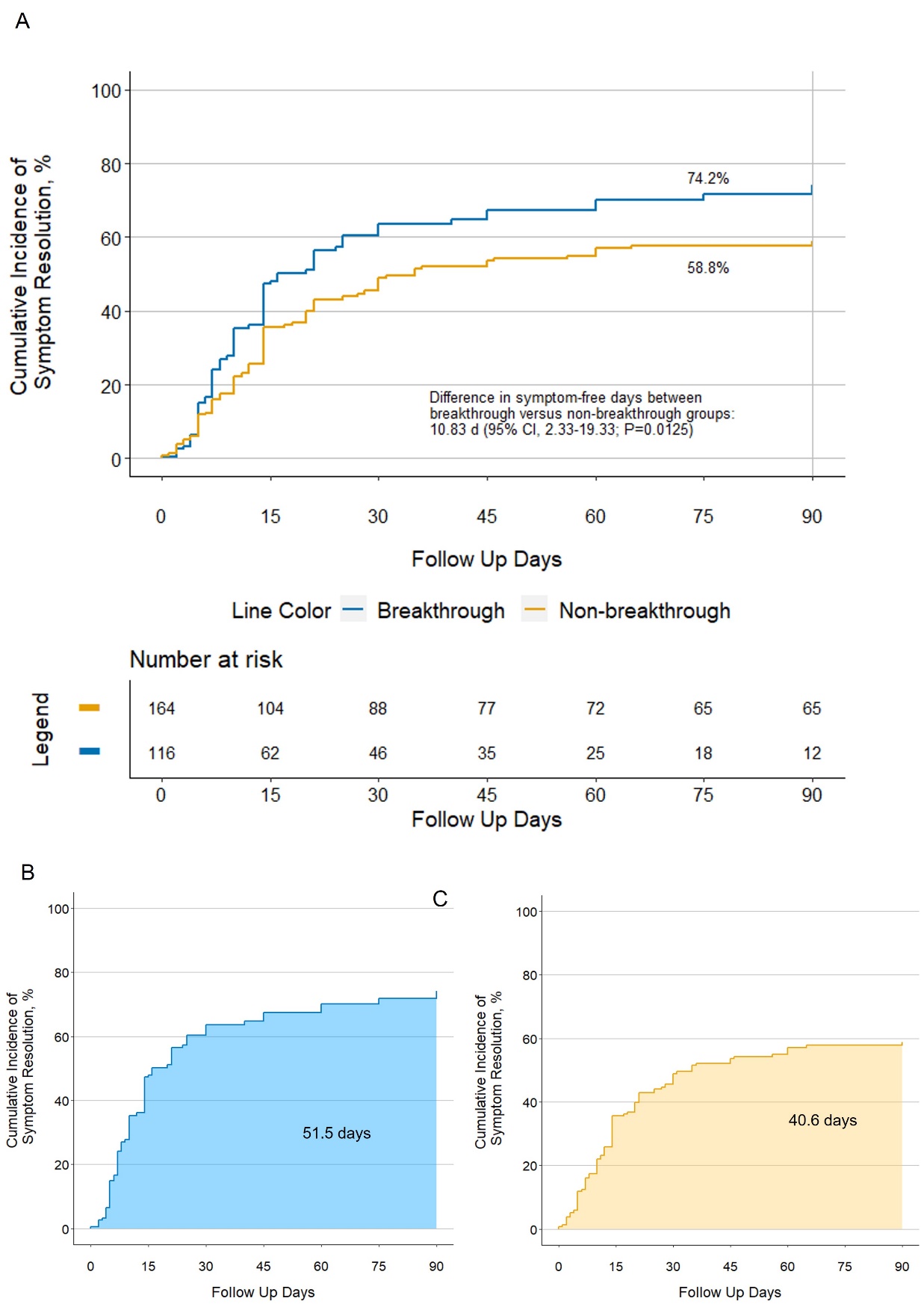

A. Cumulative incidence curves for time to symptom resolution, comparing breakthrough versus non-breakthrough infection. B and C, mean post-symptom resolution time spans as the area under the cumulative incidence curves in those with breakthrough versus non-breakthrough infection, respectively, across 90 days of follow-up.
